# GWAS of Lyme disease identifies five loci and discovers a connection with post-infectious diseases

**DOI:** 10.64898/2026.10.21.26366124

**Authors:** Satu Strausz, Erika Romppanen, Hele Haapaniemi, FinnGen, Estonian Biobank Research Team, Health Informatics Research Team, Richa Saxena, Nasa Sinnott-Armstrong, Erik Abner, Hanna M. Ollila

## Abstract

Lyme disease is a vector-borne bacterial infection with marked heterogeneity in susceptibility and clinical manifestations. We investigated the genetic architecture and clinical heterogeneity of Lyme disease using genetic and longitudinal health data from FinnGen and the Estonian Biobank. Genome-wide association analyses included 37,571 individuals with Lyme disease and 670,567 controls (N = 708,138), complemented by sex-stratified analyses and temporal phenotyping in FinnGen.

The overall GWAS meta-analysis identified three genome-wide significant loci at *SCGB1D2*, *TLR1/TLR6*, and *HLA-DQA1*. Sex-stratified analyses identified additional genome-wide significant signals at *CPE* in males (β = −0.077, P = 2.99 × 10⁻⁸) and the *TYMP-CPT1B* locus in females (β = −0.064, P = 4.98 × 10⁻⁸). The *TLR1/6* association was more pronounced in males (β = −0.141, P = 2.16 × 10⁻¹²) than in females (β = −0.060, P = 1.21 × 10⁻⁵).

Temporal phenotyping showed enrichment of neurological, musculoskeletal, and dermatological manifestations after Lyme disease diagnosis, with associations generally attenuating over time. Bell palsy showed the strongest association at 1-6 months (OR 11.49, P = 2.37 × 10⁻⁷⁹), while joint pain was also enriched during this interval (OR 1.74, P = 1.95 × 10⁻²³). Early associations with joint pain and Bell palsy were stronger in males than in females.

These findings expand the genetic architecture of Lyme disease, implicate innate immune sensing through the *TLR1/TLR6* locus, and antigen presentation through the HLA associations. Overall, the results highlight a strong polygenic signal with Lyme disease and a connection with other post-infectious clinical manifestations.

## Introduction

Lyme disease is a tick-borne infection caused by *Borrelia burgdorferi* (*Bb*) and displays marked clinical heterogeneity from early localized skin disease to disseminated involvement of the nervous system, joints, and the heart. Although most patients improve after antimicrobial therapy, a subset experience persistent or recurrent symptoms and develop post-infectious morbidity spanning multiple organ systems. The biological basis of these divergent trajectories remains incompletely understood.

Effective control of *Bb* requires rapid innate immune recognition followed by coordination of adaptive immunity. A key component of early sensing is Toll-like receptor (*TLR*) signaling, as *Bb* lipoproteins can activate inflammatory responses through *TLR2* in cooperation with *TLR1*, linking pathogen detection to downstream cytokine programs that may influence both pathogen clearance and immunopathology ^1^. These observations support a model in which host immune set points modulate susceptibility to Lyme disease and may shape the risk of downstream inflammatory sequelae.

Biological sex contributes to variation in immune function, with differences in innate and adaptive immune responses shaped by hormonal and genetic factors ^2,3,4^. These differences may influence host responses to infection, including pattern-recognition receptor signaling and downstream inflammatory pathways. This provides a rationale for examining genetic susceptibility to Lyme disease separately in females and males.

Here, we leverage large-scale biobank genetics and longitudinal health registry data from FinnGen and the Estonian Biobank to define the genetic architecture and clinical heterogeneity of Lyme disease. We perform genome-wide association analyses in the overall study population together with sex-stratified analyses to characterize shared and sex-related patterns of genetic susceptibility. We further integrate genetic findings with temporal post-diagnostic phenotyping to examine clinical manifestations following Lyme disease diagnosis and heterogeneity in disease presentation and course.

## Results

### Genome-wide association analysis implicates innate immunity

We analyzed Lyme disease liability in 708,138 individuals from FinnGen and the Estonian Biobank, including 37,571 cases and 670,567 controls (**Table 1**). Genome-wide association analysis and meta-analysis identified three loci reaching genome-wide significance (P < 5 × 10⁻⁸) (**Figure 1**, **Table 2, Supplementary Figures S1-S3**). The strongest association mapped to a missense variant annotated to *SCGB1D2* (rs2232950, β = 0.173, P = 4.45 × 10⁻¹⁰⁰), consistent with our previous report in the same biobank-based setting ^5^.

**Figure 1.**
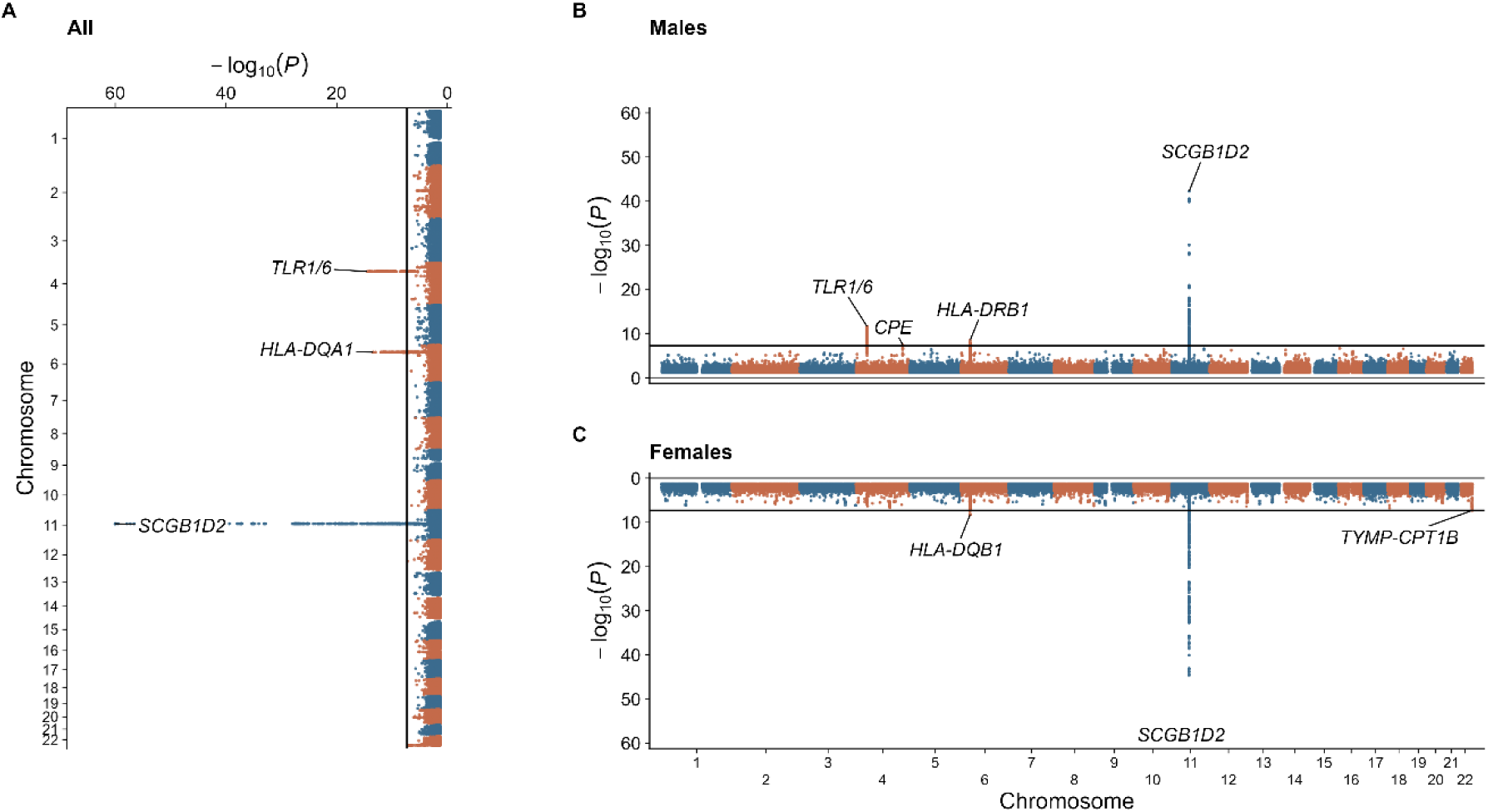
Genome-wide association analyses of Lyme disease in the overall sample and stratified by sex. Three-panel Manhattan plot showing Lyme disease association results in the full sample (A), males (B), and females (C). The black line denotes the genome-wide significance threshold (P < 5 × 10⁻⁸). For visualization, −log₁₀(P) values above 60 were truncated.

**Table 1.**
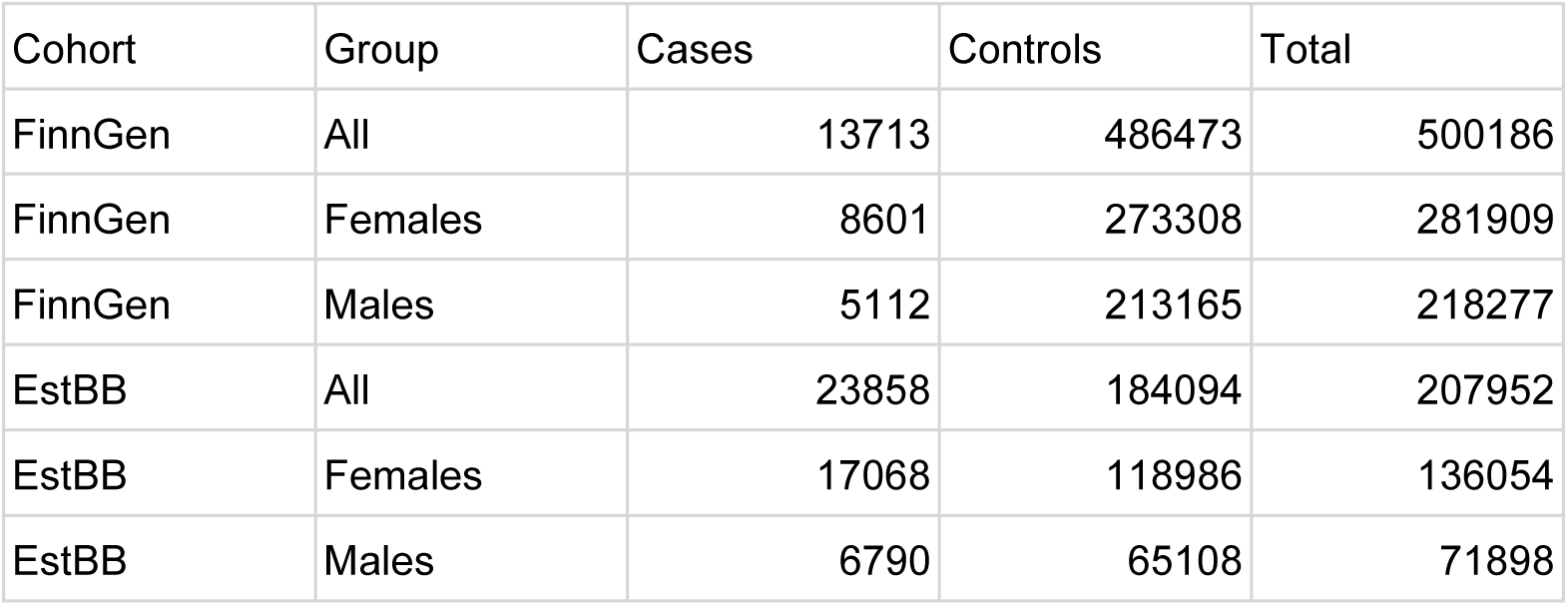
Study population. Case and control counts for Lyme disease in FinnGen and the Estonian Biobank (EstBB), overall and stratified by sex.

| Cohort | Group | Cases | Controls | Total |
| --- | --- | --- | --- | --- |
| FinnGen | All | 13713 | 486473 | 500186 |
| FinnGen | Females | 8601 | 273308 | 281909 |
| FinnGen | Males | 5112 | 213165 | 218277 |
| EstBB | All | 23858 | 184094 | 207952 |
| EstBB | Females | 17068 | 118986 | 136054 |
| EstBB | Males | 6790 | 65108 | 71898 |

**Table 2.** Lead loci associated with Lyme disease in the overall GWAS meta-analysis. Lead variants reaching genome-wide significance (P < 5×10⁻⁸) in the combined analysis. β denotes the allelic effect estimate for the alternate allele (ALT). ALT_AF indicates the alternate allele frequency. Variant annotations reflect the functional consequence for the lead variant.

| CHR | rsID | REF/ALT | ALT_AF | $\beta$ | P-value | Nearest gene(s) | Variant annotation |
| --- | --- | --- | --- | --- | --- | --- | --- |
| 4 | rs148906978 | C/T | 0.14 | -0.092 | 3.31E-15 | *TLR1/TLR6* | intronic |
| 6 | rs9271222 | C/T | 0.86 | -0.134 | 3.88E-14 | *HLA-DQA1* | intergenic |
| 11 | rs2232950 | C/T | 0.377 | 0.173 | 4.45E-100 | *SCGB1D2* | missense |

In addition, we observed genome-wide significant associations at an intronic variant in the *TLR1/6* region (rs148906978, β = −0.092, P = 3.31 × 10⁻¹⁵), within the HLA region near *HLA-DQA1* (rs9271222, β = −0.134, P = 3.88 × 10⁻¹⁴), highlighting loci implicating innate immune sensing and antigen presentation as seen with *TLR* and *HLA* genes (**Figure 1**, **Table 2**).

### Sex-stratified GWAS identifies additional Lyme disease susceptibility loci

To investigate sex-related patterns of genetic susceptibility, we performed sex-stratified GWAS separately in females and males within FinnGen and the Estonian Biobank (females: 25,669 cases and 392,294 controls; males: 11,902 cases and 278,273 controls) (Table 3, Supplementary Figures S4-S10). Among the sex-stratified results, the *TLR1/6* region showed a more pronounced association in males. The lead sex-stratified variant rs7673348 reached genome-wide significance in males (β = −0.141, P = 2.16 × 10⁻¹²), whereas the effect estimate was smaller and association evidence weaker in females (β = −0.060, P = 1.21 × 10⁻⁵) (**Table 3**). The male lead variant rs7673348 was in high linkage disequilibrium with the lead variant from the overall meta-analysis, rs148906978 (r² = 0.79, D′ = 1.00), suggesting that the two associations likely represent the same underlying *TLR1/6* signal.

**Table 3.** Sex-stratified associations for Lyme disease. Lead variants reaching genome-wide significance in either females or males, with sex-specific effect estimates. β denotes the allelic effect estimate on the log-odds scale for the alternate allele (Alt).

| Set | Locus | CHR | rsID | Ref/Alt | Alt_AF | $\beta$ female | P_female | $\beta$ male | P_male |
| --- | --- | --- | --- | --- | --- | --- | --- | --- | --- |
| Female | *HLA-DQB1* | 6 | rs12055445 | A/G | 0.514 | -5.75E-02 | **4.99E-09** | -3.67E-02 | 8.89E-03 |
| Female | *SCGB1D2* | 11 | rs2232950 | C/T | 0.376 | 1.64E-01 | **2.74E-61** | 1.95E-01 | **5.51E-43** |
| Female | *TYMP-CPT1B* | 22 | rs131788 | T/C | 0.787 | -6.38E-02 | **4.98E-08** | -1.72E-02 | 3.07E-01 |
| Male | *TLR1/TLR6* | 4 | rs7673348 | G/A | 0.145 | -6.03E-02 | 1.21E-05 | -1.41E-01 | **2.16E-12** |
| Male | *CPE* | 4 | rs9759768 | T/C | 0.485 | 4.80E-03 | 6.24E-01 | -7.65E-02 | **2.99E-08** |
| Male | *HLA-DRB1* | 6 | rs9270965 | A/G | 0.635 | -2.65E-02 | 1.05E-02 | -8.72E-02 | **2.91E-09** |
| Male | *SCGB1D2* | 11 | rs2232950 | C/T | 0.379 | 0.164 | **2.74E-61** | 0.195 | **5.51E-43** |

Outside the *TLR1/6* region, we observed additional sex-related signals. In males, we detected a genome-wide significant association at *CPE* (rs9759768, β = −0.077, P = 2.99 × 10⁻⁸), with little evidence of association in females (β = 0.005, P = 0.62) (Table 3). In females, we observed a genome-wide significant association at the *TYMP-CPT1B* locus on chromosome 22 (rs131788, β = −0.064, P = 4.98 × 10⁻⁸), whereas the corresponding estimate was smaller in males (β = −0.017, P = 0.31) (Table 3).

### Shared and sex-related signals across the HLA region and non-HLA loci

Sex-stratified analyses also showed different association patterns across the HLA region, with a female signal near *HLA-DQB1* (rs12055445, P_female = 4.99 × 10⁻⁹; P_male = 8.89 × 10⁻³) and a male signal near *HLA-DRB1* (rs9270965, P_male = 2.91 × 10⁻⁹; P_female = 0.01) (Table 3). The two lead variants were in low linkage disequilibrium (r² = 0.05). In contrast, *SCGB1D2* showed strong association evidence in both females and males (P_female = 2.73 × 10⁻⁶¹; P_male = 5.51 × 10⁻⁴³). These findings suggest that Lyme disease susceptibility includes both broadly shared genetic associations and loci with sex-related differences in association strength.

### Sex-stratified PheWAS of the *TLR1/6* lead variant

In sex-stratified PheWAS of the *TLR1/6* lead variant rs148906978, the strongest non-Lyme association in females was observed with allergic rhinitis (β = −0.149, P = 3.09 × 10⁻¹⁰). In males, the strongest non-Lyme association was observed with a childhood asthma phenotype (β = −0.195, P = 1.09 × 10⁻⁵), with additional nominal associations with allergic asthma, allergic rhinitis, and several infection-related phenotypes in both sexes (Supplementary Figure 11).

### Temporal patterns of clinical manifestations following Lyme disease diagnosis

To characterize clinical trajectories following Lyme disease diagnosis, we performed temporal phenome-wide analyses of any diagnosis or medication with post-diagnostic diagnoses using electronic health record data in FinnGen and 2,001 diagnoses. Several neurological, musculoskeletal, and dermatological manifestations remained enriched beyond the acute diagnostic period (**Figure 2, Supplementary Table S1**). Bell palsy showed the strongest association at 1-6 months after diagnosis (OR 11.49, P = 2.37 × 10⁻⁷⁹), with progressive attenuation at 6-12 months (OR 4.52, P = 3.18 × 10⁻¹⁵) and 1-2 years (OR 2.35, P = 4.26 × 10⁻⁵). Paraesthesia of the skin similarly remained enriched across all three intervals (OR 2.87, P = 6.81 × 10⁻¹⁵; OR 2.77, P = 3.20 × 10⁻¹⁴; and OR 1.79, P = 3.01 × 10⁻⁶, respectively). Associations were also observed across all three intervals for joint pain (OR 1.74, P = 1.95 × 10⁻²³; OR 1.58, P = 2.08 × 10⁻¹⁶; and OR 1.42, P = 1.84 × 10⁻¹⁴) and dermatitis (OR 2.20, P = 4.05 × 10⁻³²; OR 1.68, P = 3.68 × 10⁻¹³; and OR 1.51, P = 5.09 × 10⁻¹²).

**Figure 2.**
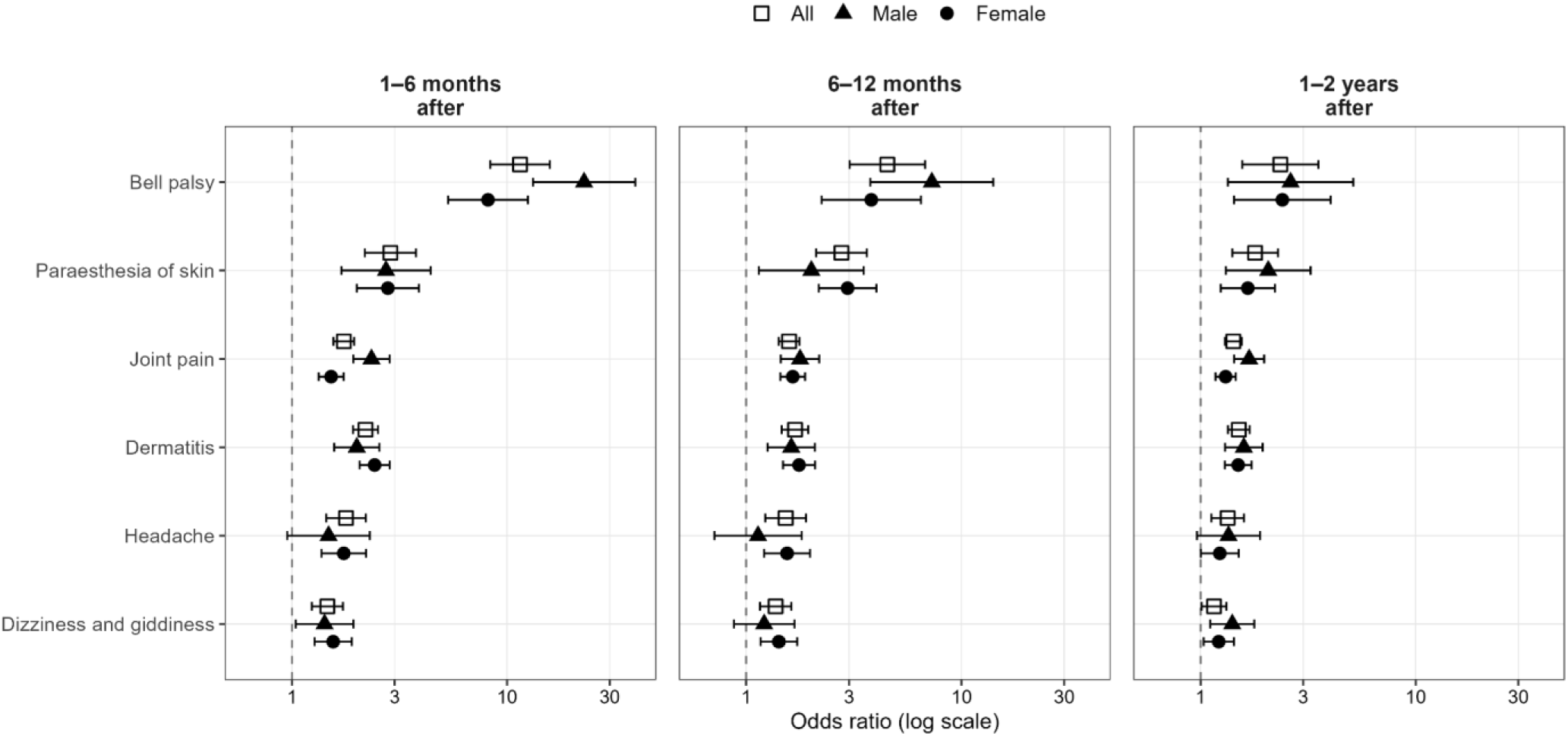
Post-diagnostic clinical manifestations following Lyme disease diagnosis. Odds ratios (ORs) and 95% confidence intervals are shown for neurological, musculoskeletal, and dermatological manifestations at 1-6 months, 6-12 months, and 1-2 years after Lyme disease diagnosis. Estimates are presented for the overall study population and separately for males and females. The vertical dashed line indicates OR = 1.

Headache and dizziness showed more modest associations that attenuated over time; for dizziness, ORs were 1.46 (P = 9.32 × 10⁻⁶), 1.37 (P = 2.57 × 10⁻⁴), and 1.15 (P = 0.037) across the three post-diagnostic intervals.

Sex-stratified analyses showed broadly concordant trajectories in females and males, with some stronger early post-diagnostic associations in males (Figure 2, **Supplementary Table S2**). At 1– 6 months, joint pain was more strongly associated in males (OR 2.34, P = 1.85 × 10⁻¹⁸) than in females (OR 1.52, P = 8.36 × 10⁻¹⁰; P_sex = 3.35 × 10⁻⁴, FDR q = 0.006). Bell palsy showed a larger difference in effect size, with ORs of 22.80 in males (P = 2.29 × 10⁻⁵⁸) and 8.15 in females (P = 1.25 × 10⁻²⁹; P_sex = 0.0037, FDR q = 0.033).

### Cross-phenotype associations of Lyme disease lead variants

Cross-phenotype look-up of the overall Lyme disease GWAS lead variants in FinnGen identified nominal associations (P < 0.05) with several phenotypes corresponding to or related to the clinical manifestations highlighted by the temporal analysis (**Supplementary Table S3**). The *SCGB1D2* lead variant rs2232950 was associated with joint pain, Bell palsy, and dermatitis-related phenotypes. The *HLA-DQA1* region variant showed strong associations with dermatitis-related phenotypes, together with nominal associations with joint pain and Bell palsy (**Supplementary Table 3**). No nominal associations with the focused clinical phenotypes were observed for the *TLR1/6* lead variant.

## Discussion

### Overview

In this biobank-scale study of Lyme disease susceptibility across FinnGen and the Estonian Biobank, we identified genome-wide significant loci implicating innate immune sensing and antigen presentation, alongside both shared and sex-related patterns of genetic association. The overall meta-analysis identified associations at *TLR1/6*, *HLA-DQA1* and *SCGB1D2* while sex-stratified analyses identified additional signals at *CPE* in males and the *TYMP-CPT1B* locus in females. The *TLR1/6* association was stronger in males, whereas *SCGB1D2* showed broadly consistent effects across sexes. Temporal phenotyping further identified neurological, musculoskeletal, and dermatological manifestations following Lyme disease diagnosis, with joint pain and Bell palsy showing stronger early post-diagnostic associations in males. These findings highlight heterogeneity in both genetic susceptibility and clinical manifestations of Lyme disease.

### Innate immune sensing and sex-related patterns at the *TLR1/6* region

The association at the *TLR1/6* region is biologically plausible given the central role of Toll-like receptor signaling in the recognition of *Borrelia burgdorferi*. *Borrelia* lipoproteins can activate inflammatory responses through *TLR2*, with *TLR1/TLR2* heterodimers playing an important role in the early human inflammatory response to *Borrelia* ^6^. Previous studies of the *TLR1* p.Ser602Ile polymorphism (rs5743618) have linked variation at this site to heightened inflammatory responses and antibiotic-refractory Lyme arthritis ^7^, while more recent functional work suggests a role in impaired innate immune tolerance following *B. burgdorferi* stimulation ^8^. This missense variant is in linkage disequilibrium with our overall lead variant rs148906978 (r² = 0.57), linking our association signal to previously described functional variation in *TLR1*.

In our sex-stratified analyses, the *TLR1/6* association was more pronounced in males than in females. Previous studies have reported sex-related differences in cytokine responses associated with Lyme borreliosis ^9^, hematological responses during acute untreated disease ^10^, and, more recently, serologic response and severity of early Lyme disease ^11^. Our sex-stratified PheWAS identified associations beyond Lyme disease, including a strong association with allergic rhinitis in females and associations with asthma-related phenotypes in both sexes. Nominal associations with other infection-related phenotypes were also observed in both sexes.

More broadly, sex-related differences in immunity are shaped by hormonal and genetic mechanisms that influence innate and adaptive immune responses ^2,12,13^. Such differences in immune state may contribute to variation in how inherited changes in pathogen-sensing pathways are manifested clinically. The biological basis of the stronger *TLR1/6* association observed in males remains unclear, and functional studies integrating genotype with immune phenotypes will be needed to clarify how variation at this locus influences Lyme disease susceptibility in females and males.

### Shared and sex-related contributions across HLA and non-HLA loci

Beyond innate immune sensing, our results implicate both shared and sex-related contributions from other susceptibility loci. The HLA region was associated with Lyme disease in the overall analysis, while sex-stratified analyses highlighted different lead signals near *HLA-DQB1* in females and *HLA-DRB1* in males. Previous studies have implicated HLA class II variation in Lyme disease, particularly in Lyme arthritis, including associations of *HLA-DRB1-DQA1-DQB1* haplotypes carrying *DRB1*04* or *DRB1*01* alleles with antibiotic-refractory disease ^5,14^. Given the central role of HLA class II molecules in antigen presentation, variation in this region may influence the host response to *Borrelia* through differences in antigen recognition and downstream adaptive immune activation. However, because of the extensive linkage disequilibrium and genetic complexity of the HLA region, differences in lead variants between the sex-stratified analyses should not necessarily be interpreted as distinct causal effects without further functional characterization.

In contrast, the previously reported missense association in *SCGB1D2* showed broadly similar effects in females and males. In our previous study, the *SCGB1D2* P53L variant was associated with Lyme disease susceptibility, and functional experiments demonstrated that SCGB1D2 inhibits *B. burgdorferi* growth in vitro and limits establishment of infection in vivo ^5^. Sex-stratified analyses additionally identified genome-wide significant signals at *CPE* in males and the *TYMP-CPT1B* locus in females. The *CPE* lead variant also showed a nominal association with nummular dermatitis in FinnGen. As nummular dermatitis can resemble erythema migrans, diagnostic overlap could contribute to this cross-phenotype association ^15^. The *TYMP-CPT1B* locus has also been associated with neurological autoimmune diseases, most notably narcolepsy ^16^. The results suggest a genetic architecture comprising both broadly shared susceptibility loci and selected loci with more pronounced sex-related patterns.

### Sex differences in post-diagnostic clinical manifestations

Our temporal phenotyping complements the genetic findings by showing that neurological, musculoskeletal, and dermatological manifestations remain enriched after Lyme disease diagnosis, with several associations attenuating over time. Persistent pain, fatigue, and neurological or cognitive symptoms have previously been described following treated Lyme disease, although attribution of these symptoms to the preceding infection remains challenging because of their substantial background prevalence ^17,18,19^.

Notably, joint pain and Bell palsy showed stronger associations in males during the 1-6 month interval. Previous studies suggest that sex differences in Lyme disease vary by age and clinical manifestation. In a US study of more than 88,000 patients, disseminated manifestations were more common in males, with particularly increased risks of Lyme arthritis and facial palsy in younger males ^20^. A recent prospective study similarly reported greater early disease severity and higher seropositivity in males, particularly compared with premenopausal females ^11^. In contrast, long-term follow-up of culture-confirmed early Lyme disease found no appreciable sex differences in post-treatment symptoms ^21^. These observations suggest that sex-related differences may be most apparent for specific manifestations and phases of Lyme disease rather than representing a uniform difference in disease course.

### Limitations

This study should be interpreted in the light of the following limitations. First, Lyme disease case status was based on registry and EHR-derived phenotypes and therefore captures clinically diagnosed disease rather than uniformly laboratory-confirmed infection; non-differential misclassification and possible underdiagnosis would be expected to attenuate effect estimates. Second, Lyme disease risk reflects both pathogen exposure and host response, and sex differences in exposure patterns, healthcare utilization, or diagnostic pathways may contribute to the observed differences between females and males. Third, sex-stratified analyses reduce effective sample size, limiting power to detect more modest associations beyond the strongest loci. Finally, differences in lead variants between the overall and sex-stratified analyses may reflect differences in statistical power and sampling variation among correlated variants within associated loci; additional validation will be needed to pinpoint causal variants and effector genes.

### Conclusions

In summary, our results characterize the genetic architecture of Lyme disease susceptibility in two large European biobanks and implicate host pathways involved in innate immune sensing and antigen presentation. We identify both broadly shared and sex-related genetic associations, alongside differences in selected post-diagnostic clinical manifestations between females and males. These findings highlight heterogeneity in Lyme disease susceptibility and clinical presentation and provide a basis for further studies of the host mechanisms contributing to variation in disease risk and course.

## Materials and Methods

### Cohort description and study population

FinnGen is a nationwide public-private partnership combining genotype data from Finnish biobank participants with longitudinal outcomes from national health registers. Genotyping, sample- and variant-level quality control, and genotype imputation were performed within FinnGen using standard pipelines as described previously ^22^. In this study, we analysed 500,186 FinnGen participants (13,713 Lyme disease cases; 486,473 controls), including 281,909 females (8,601 cases; 273,308 controls) and 218,277 males (5,112 cases; 213,165 controls).

The Estonian Biobank is a volunteer-based cohort maintained by the Institute of Genomics, University of Tartu, with broad consent and linkage to national electronic health records and registries enabling longitudinal phenotyping ^23,24^. Genotyping, quality control, and imputation were conducted within EstBB using established biobank pipelines as described previously ^5^. In this study, we analysed 207,952 EstBB participants (23,858 cases; 184,094 controls), including 136,054 females (17,068 cases; 118,986 controls) and 71,898 males (6,790 cases; 65,108 controls).

### Genome-wide association analyses

We conducted genome-wide association analyses for Lyme disease case-control status separately in FinnGen and EstBB using REGENIE ^25^. Association testing used an additive genetic model with imputed genotype dosages and included age, sex (in non-stratified analyses), genotyping batch, and genetic principal components as covariates. For unbalanced case-control settings, we used REGENIE’s robust option for binary traits (approximate Firth correction as implemented in REGENIE) to improve calibration.

We performed sex-stratified GWAS in females and males within each cohort using the same framework and covariates (excluding sex). Genome-wide significance was defined as P < 5×10⁻⁸.

### Meta-analysis

We combined FinnGen and EstBB summary statistics using fixed-effects inverse-variance weighted meta-analysis implemented in METAL ^26^. Meta-analyses were performed for the overall GWAS as well as for sex-stratified GWAS, and heterogeneity across cohorts was assessed using Cochran’s Q test.

### Temporal phenotyping

To characterize clinical manifestations around and following Lyme disease diagnosis, we performed temporal code-wide association analyses (TimeCodeWAS). For each case cohort, up to 10 controls were matched by sex and year of birth. Time was defined relative to the Lyme disease diagnosis date in cases and the corresponding index date in matched controls. Clinical diagnoses were evaluated in three non-overlapping post-diagnostic time windows: 1-6 months, 6-12 months, and 1-2 years after the index date. TimeCodeWAS compared the occurrence of condition-based medical codes between Lyme disease cases and matched controls within each post-diagnostic time window. Code enrichment was evaluated using chi-square tests and summarized as odds ratios (ORs). To account for multiple testing in this focused analysis, Benjamini-Hochberg false discovery rate (FDR) correction was applied jointly across the 18 sex-difference tests comprising six clinical manifestations and three post-diagnostic time windows.

## Ethics statement

Study subjects in FinnGen provided informed consent for biobank research, based on the Finnish Biobank Act. Alternatively, separate research cohorts, collected prior the Finnish Biobank Act came into effect (in September 2013) and start of FinnGen (August 2017), were collected based on study-specific consents and later transferred to the Finnish biobanks after approval by Fimea (Finnish Medicines Agency), the National Supervisory Authority for Welfare and Health. Recruitment protocols followed the biobank protocols approved by Fimea. The Coordinating Ethics Committee of the Hospital District of Helsinki and Uusimaa (HUS) statement number for the FinnGen study is Nr HUS/990/2017.

The FinnGen study is approved by Finnish Institute for Health and Welfare (permit numbers: THL/2031/6.02.00/2017, THL/1101/5.05.00/2017, THL/341/6.02.00/2018, THL/2222/6.02.00/2018, THL/283/6.02.00/2019, THL/1721/5.05.00/2019 and THL/1524/5.05.00/2020), Digital and population data service agency (permit numbers: VRK43431/2017-3, VRK/6909/2018-3, VRK/4415/2019-3), the Social Insurance Institution (permit numbers: KELA 58/522/2017, KELA 131/522/2018, KELA 70/522/2019, KELA 98/522/2019, KELA 134/522/2019, KELA 138/522/2019, KELA 2/522/2020, KELA 16/522/2020), Findata permit numbers THL/2364/14.02/2020, THL/4055/14.06.00/2020, THL/3433/14.06.00/2020, THL/4432/14.06/2020, THL/5189/14.06/2020, THL/5894/14.06.00/2020, THL/6619/14.06.00/2020, THL/209/14.06.00/2021, THL/688/14.06.00/2021, THL/1284/14.06.00/2021, THL/1965/14.06.00/2021, THL/5546/14.02.00/2020, THL/2658/14.06.00/2021, THL/4235/14.06.00/2021, THL/4990/14.02.00/2023 Statistics Finland (permit numbers: TK-53-1041-17 and TK/143/07.03.00/2020 (earlier TK-53-90-20) TK/1735/07.03.00/2021, TK/3112/07.03.00/2021) and Finnish Registry for Kidney Diseases permission/extract from the meeting minutes on 4th July 2019.

The Biobank Access Decisions for FinnGen samples and data utilized in FinnGen Data Freeze 13 include: THL Biobank BB2017_55, BB2017_111, BB2018_19, BB_2018_34, BB_2018_67, BB2018_71, BB2019_7, BB2019_8, BB2019_26, BB2020_1, BB2021_65, BB22-0025-A01, BB22-0025-A03, BB23-0222-A01, BB22-0025-A04, BB22-0025-A06, BB22-0025-A08, THLBB2024_30. Finnish Red Cross Blood Service Biobank 7.12.2017, 13.11.2023, 001-2023, Helsinki Biobank HUS/359/2017, HUS/248/2020, HUS/430/2021 §28, §29, HUS/150/2022 §12, §13, §14, §15, §16, §17, §18, §23, §58, §59, HUS/128/2023 §18, BB22-0025-A01, BB22-0025-A02, BB22-0025-A05, BB22-0025-A07, BB22-0025-A09, BB22-0025-A10, BB22-0025-A03, BB23-0222-A01, BB22-0025-A04, BB22-0025-A06, BB22-0025-A08, Amendment_BB22-0025-A05, Decision allowing to continue data processing until 31st Aug 2027: BB_2021-0140, HUS/150/2022 §12, BB_2021-0139, HUS/150/2022 §13, BB_2021-0161,HUS/150/2022 §14, BB_2021-0164, HUS/150/2022 §15, BB_2021-0169, HUS/150/2022 §16, BB_2021-0170, HUS/150/2022 §17, BB_2021-0179, HUS/150/2022 §18, BB_2022-0262, HUS/150/2022 §58, BB22-0067, HUS/150/2022 §59, Auria Biobank AB17-5154 and amendment #1 (August 17 2020) and amendments BB_2021-0140, BB_2021-0156 (August 26 2021, Feb 2 2022), BB_2021-0169, BB_2021-0179, BB_2021-0161, AB20-5926 and amendment #1 (April 23 2020) and it’s modifications (Sep 22 2021), BB_2022-0262, BB_2022-0256, BB22-0025-A01, BB22-0025-A02, BB22-0025-A03, BB23-0222_A01, BB22-0025-A02, BB22-0025-A05, BB22-0025-A07, BB22-0025-A09, BB22-0025-A10, BB22-0025-A03, BB23-0222-A01, BB22-0025-A04, BB22-0025-A06, BB22-0025-A08, Decision allowing to continue data processing until 31st Aug 2027: AB20-5926, BB_2021-0140, BB_2021-0156, BB_2021-0161, BB_2021-0161, BB_2021-0164, BB_2021-0169, BB_2021-0179, BB_2022-0262, Biobank Borealis of Northern Finland_2017_1013, 2021_5010, 2021_5010 Amendment, 2021_5018, 2021_5018 Amendment, 2021_5015, 2021_5015 Amendment, 2021_5015 Amendment_2, 2021_5023, 2021_5023 Amendment, 2021_5023 Amendment_2, 2021_5017, 2021_5017 Amendment, 2022_6001, 2022_6001 Amendment, 2022_6006 Amendment, 2022_6006 Amendment_2, BB22-0067, 2022_0262, 2022_0262 Amendment, BB22-0025-A01, BB22-0025-A02, BB22-0025-A05, BB22-0025-A07, BB22-0025-A09, BB22-0025-A10, BB22-0025-A03, BB23-0222-A01, BB22-0025-A04, BB22-0025-A06, BB22-0025-A08, Decision allowing to continue data processing until 31st Aug 2027: BB/2021/5015, BB/2021/5017, BB/2021/5018, BB/2021/5023, BB/2022/6006, BB/2022/6001, BB/2022-0262, BB/2021/5010, Biobank of Eastern Finland 1186/2018 and amendment 22§/2020, 53§/2021, 13§/2022, 14§/2022, 15§/2022, 27§/2022, 28§/2022, 29§/2022, 33§/2022, 35§/2022, 36§/2022, 37§/2022, 39§/2022, 7§/2023, 32§/2023, 33§/2023, 34§/2023, 35§/2023, 36§/2023, 37§/2023, 38§/2023, 39§/2023, 40§/2023, 41§/2023, BB22-0025-A01, BB22-0025-A02, BB22-0025-A05, BB22-0025-A07, BB22-0025-A09, BB22-0025-A10, BB22-0025-A03, BB23-0222-A01, BB22-0025-A04, BB22-0025-A06, BB22-0025-A08, Decision allowing to continue data processing until 31st Aug 2027: MO-BB_2021-0179-A0, MO-BB_2021-0156_PRE-A01, BB_2021-0140, MO-BB_2021-0170_PRE-A0, MO-BB_2021-0169-A01, MO-BB_2022-0256-A01, MO-BB_2021-0161-A01, MO-BB_2021-0161-A02, BB22-0067-A01, MO-BB_2022-0262-A0, Finnish Clinical Biobank Tampere MH0004 and amendments (21.02.2020 & 06.10.2020), BB2021-0140 8§/2021, 9§/2021, §9/2022, §10/2022, §12/2022, 13§/2022, §20/2022, §21/2022, §22/2022, §23/2022, 28§/2022, 29§/2022, 30§/2022, 31§/2022, 32§/2022, 38§/2022, 40§/2022, 42§/2022, 1§/2023, BB2021-0140, BB22-0025-A01, BB_2021-0161, BB22-0025-A02, BB22-0025-A05, BB22-0025-A07, BB22-0025-A09, BB22-0025-A10, BB22-0025-A03, BB23-0222-A01, BB22-0025-A04, BB22-0025-A06, BB22-0025-A08, Decision allowing to continue data processing until 31st Aug 2027: BB_2021-0140, BB_ 2021-0161, BB_ 2021-0179, BB_ 2021-0156, BB_ 2021-0169, BB_ 2021-0170, BB22-0067-A01, Central Finland Biobank 1-2017, BB_2021-0169, BB_2021-0179, BB_2022-0256, BB_2022-0262, Decision allowing to continue data processing until 31st Aug 2027 for projects: BB_2021-0179, BB22-0067,BB_2022-0262, BB_2021-0170, BB_2021-0164, BB_2021-0161, and BB_2021-0169, BB22-0025-A01, BB22-0025-A02, BB22-0025-A05, BB22-0025-A07, BB22-0025-A09, BB22-0025-A10, BB22-0025-A03, BB23-0222-A01, BB22-0025-A04, BB22-0025-A06, BB22-0025-A08, Terveystalo Biobank STB 2018001 and amendment 25th Aug 2020, Finnish Hematological Registry and Clinical Biobank decision 18th June 2021, Amendment 2nd January 2024 and Arctic biobank P0844: ARC_2021_1001, ARC_2023_3003 (BB22-0025-A01), BB22-0025-A03, BB23-0222-A01, BB22-0025-A04, BB22-0025-A06, BB22-0025-A08.

The Estonian Biobank is a volunteer-based biobank with 212,948 participants in the current data freeze (2026v03). All biobank participants have signed a broad informed consent form and information on ICD-10 codes is obtained via regular linking with the national Health Insurance Fund and other relevant databases, with majority of the electronic health records having been collected since 2004. Analyses were restricted to individuals with European ancestry. The activities of the EstBB are regulated by the Human Genes Research Act, which was adopted in 2000 specifically for the operations of EstBB. Individual level data analysis in EstBB was carried out under ethical approval 1.1-12/624 from the Estonian Committee on Bioethics and Human Research (Estonian Ministry of Social Affairs), using data according to release application 6-7/GI/24953 from the Estonian Biobank.

## Data availability

Genome-wide association summary statistics will be made available through the NHGRI-EBI GWAS Catalog under accession number XXX upon publication. Individual-level genotype and registry data are not publicly available because of participant consent and data protection requirements but may be accessed through the respective FinnGen and Estonian Biobank data access procedures. FinnGen data can be accessed through the Fingenious® services, and Estonian Biobank data through an approved data access application.

## Acknowledgements

We want to acknowledge the participants and investigators of the FinnGen study. We want to acknowledge the participants of the Estonian Biobank for their contributions. The Estonian Genome Center analyses were partially carried out in the High Performance Computing Center, University of Tartu. The Estonian Biobank Research Team was responsible for data collection, genotyping, quality control and imputation and consisted of Andres Metspalu, Mait Metspalu, Lili Milani, Reedik Mägi, Mari Nelis, Tõnu Esko and Georgi Hudjashov.

