## Supplementary_Figures for "GWAS of Lyme disease identifies five loci and discovers a connection with post-infectious diseases"

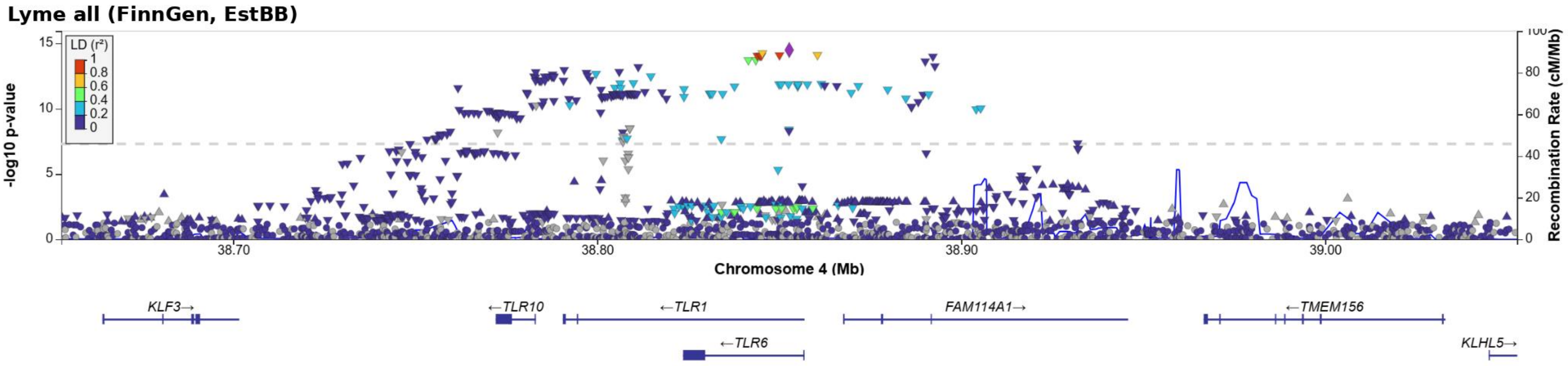


**Supplementary Figure S1.** Regional association plot for the *TLR1/TLR6* locus (chromosome 4) in the overall Lyme disease GWAS meta-analysis. Points are colored according to linkage disequilibrium (LD, $r^2$) with the lead variant. The dashed horizontal line indicates genome-wide significance ($P = 5 \times 10^{-8}$). The blue line denotes recombination rate, and gene annotations are shown below the association plot.

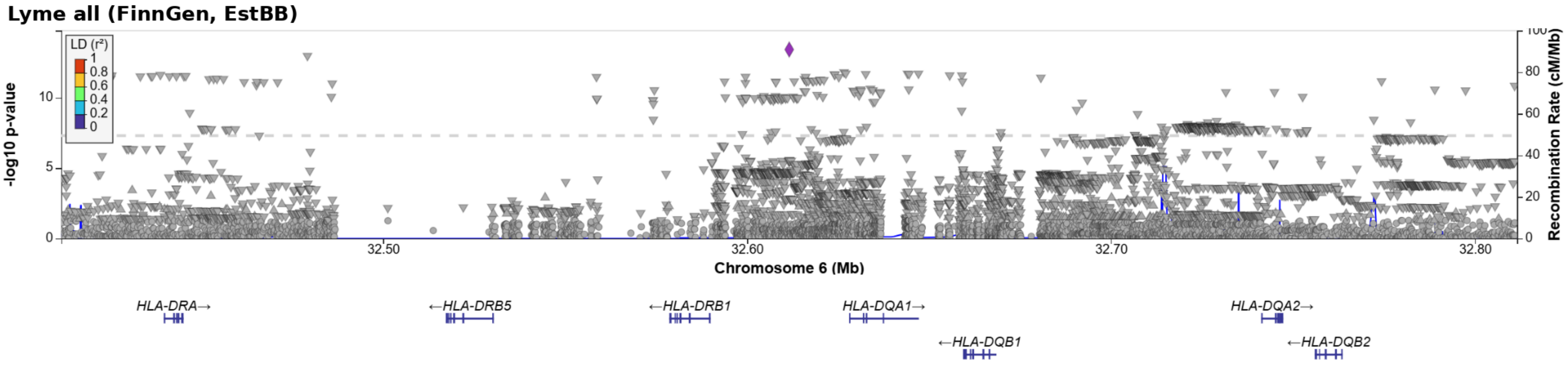


**Supplementary Figure S2.** Regional association plot for the HLA locus (chromosome 6) in the overall Lyme disease GWAS meta-analysis. Points are colored according to linkage disequilibrium (LD, $r^2$) with the lead variant. The dashed horizontal line indicates genome-wide significance ($P = 5 \times 10^{-8}$). The blue line denotes recombination rate, and gene annotations are shown below the association plot.

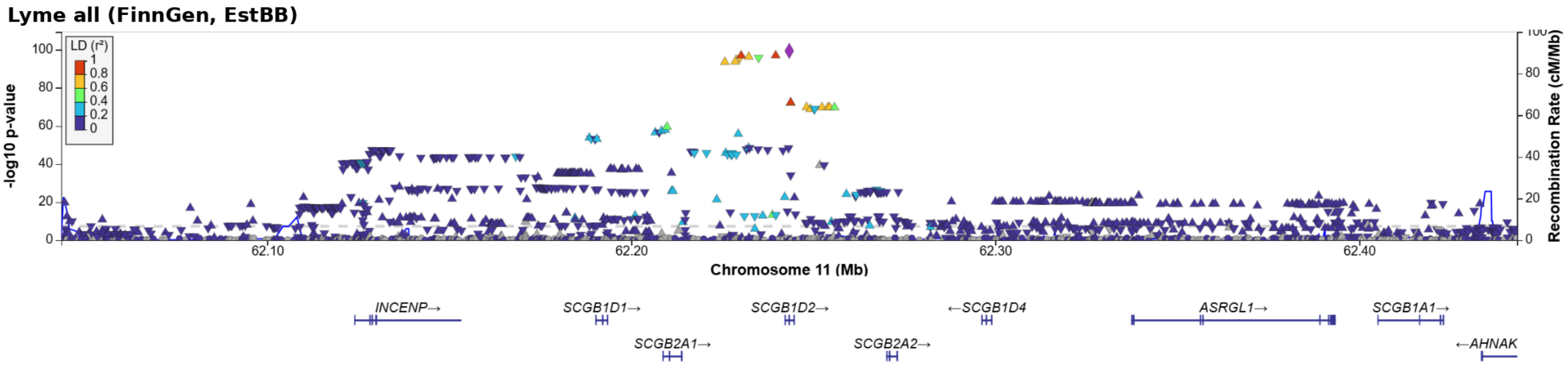


**Supplementary Figure S3.** Regional association plot for the *SCGB1D2* locus (chromosome 11) in the overall Lyme disease GWAS meta-analysis. Points are colored according to linkage disequilibrium (LD, $r^2$) with the lead variant. The dashed horizontal line indicates genome-wide significance ($P = 5 \times 10^{-8}$). The blue line denotes recombination rate, and gene annotations are shown below the association plot.

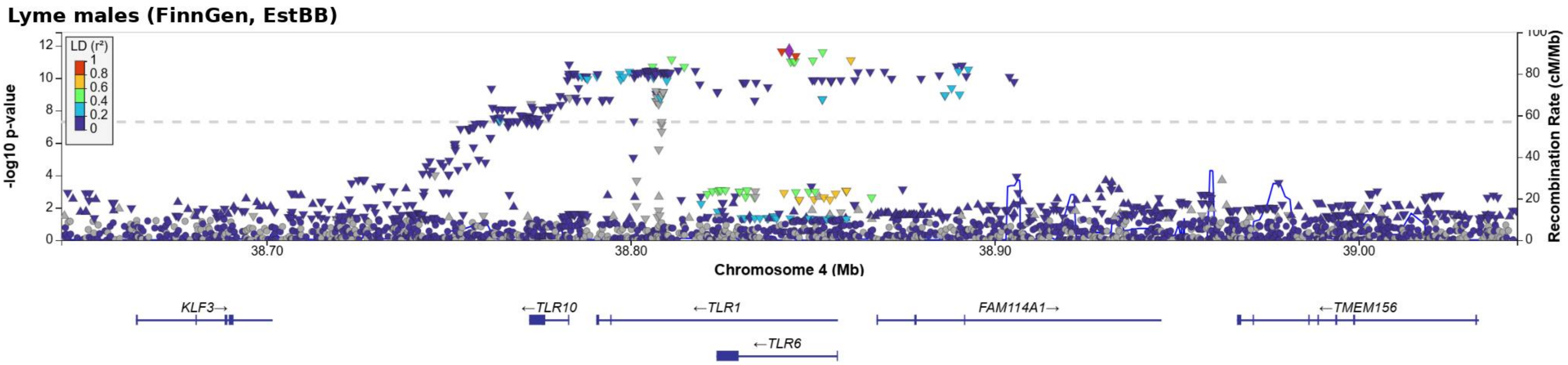


**Supplementary Figure S4.** Regional association plot for the *TLR1/TLR6* locus (chromosome 4) in the male-stratified Lyme disease GWAS meta-analysis. Points are colored according to linkage disequilibrium (LD, $r^2$) with the lead variant. The dashed horizontal line indicates genome-wide significance ($P = 5 \times 10^{-8}$). The blue line denotes recombination rate, and gene annotations are shown below the association plot.

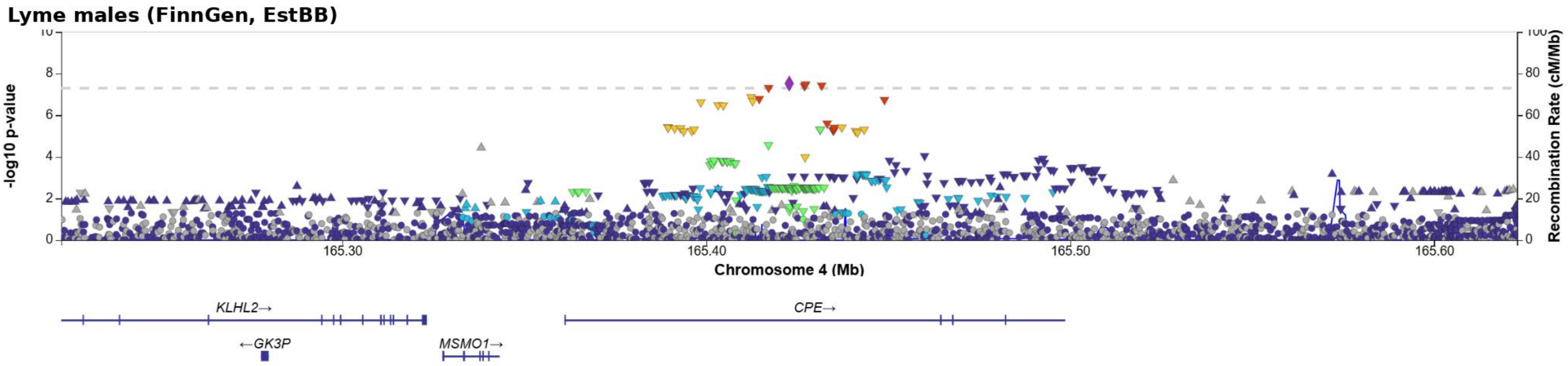


**Supplementary Figure S5.** Regional association plot for the *CPE* locus (chromosome 4) in the male-stratified Lyme disease GWAS meta-analysis. Points are colored according to linkage disequilibrium (LD, $r^2$) with the lead variant. The dashed horizontal line indicates genome-wide significance ($P = 5 \times 10^{-8}$). The blue line denotes recombination rate, and gene annotations are shown below the association plot.

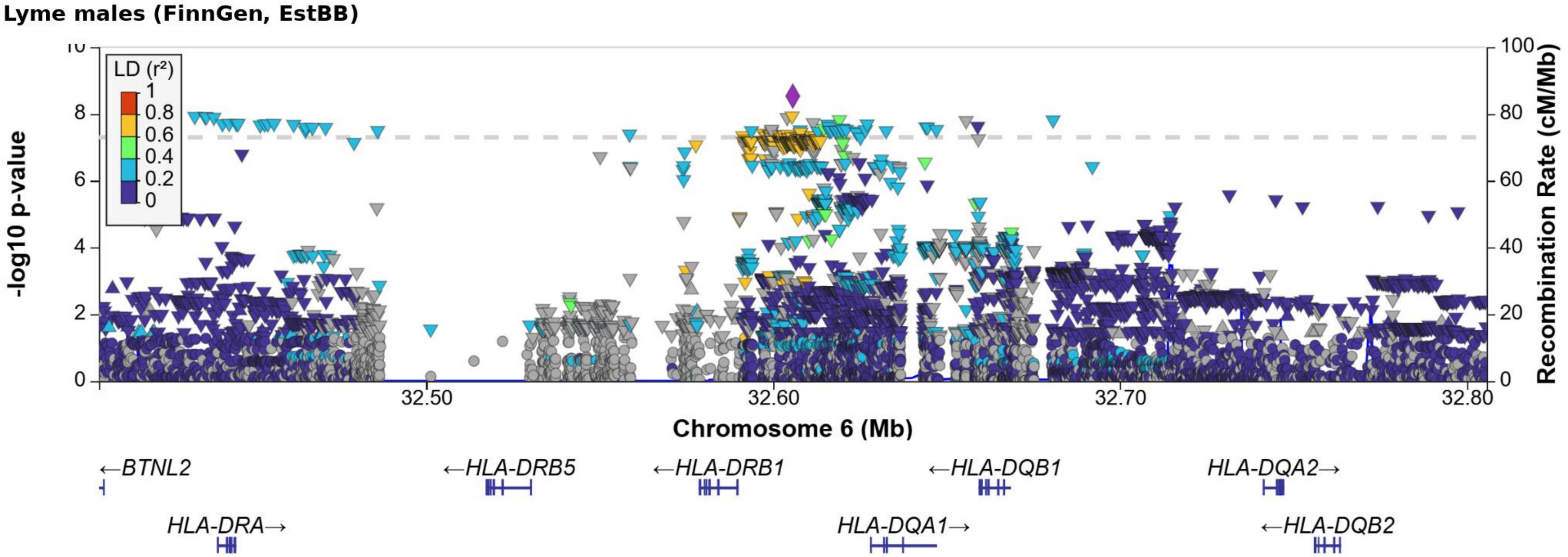


**Supplementary Figure S6.** Regional association plot for the HLA locus (chromosome 6) in the male-stratified Lyme disease GWAS meta-analysis. Points are colored according to linkage disequilibrium (LD, $r^2$) with the lead variant. The dashed horizontal line indicates genome-wide significance ($P = 5 \times 10^{-8}$). The blue line denotes recombination rate, and gene annotations are shown below the association plot.

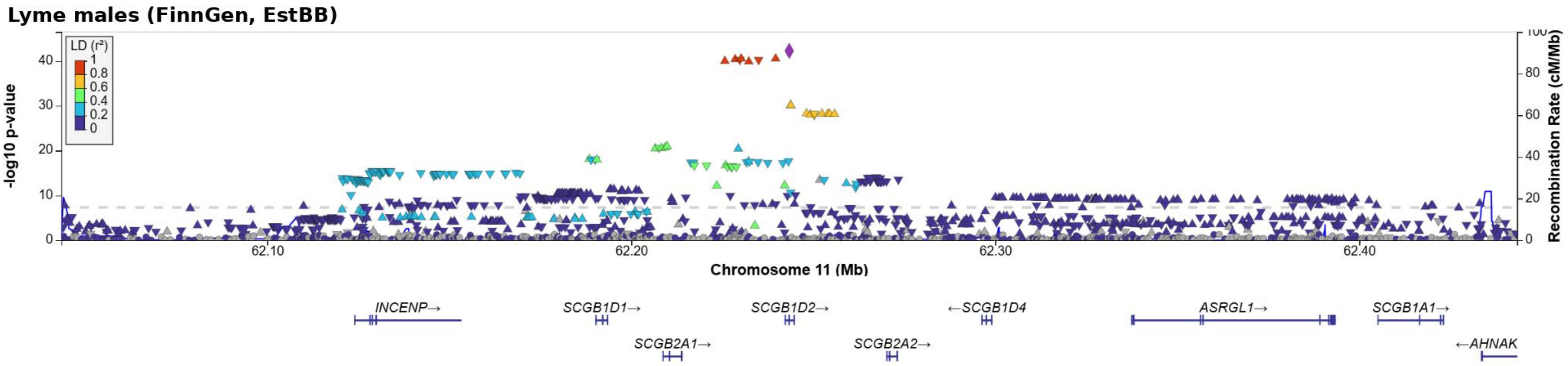


**Supplementary Figure S7.** Regional association plot for the *SCGB1D2* locus (chromosome 11) in the male-stratified Lyme disease GWAS meta-analysis. Points are colored according to linkage disequilibrium (LD, $r^2$) with the lead variant. The dashed horizontal line indicates genome-wide significance ($P = 5 \times 10^{-8}$). The blue line denotes recombination rate, and gene annotations are shown below the association plot.

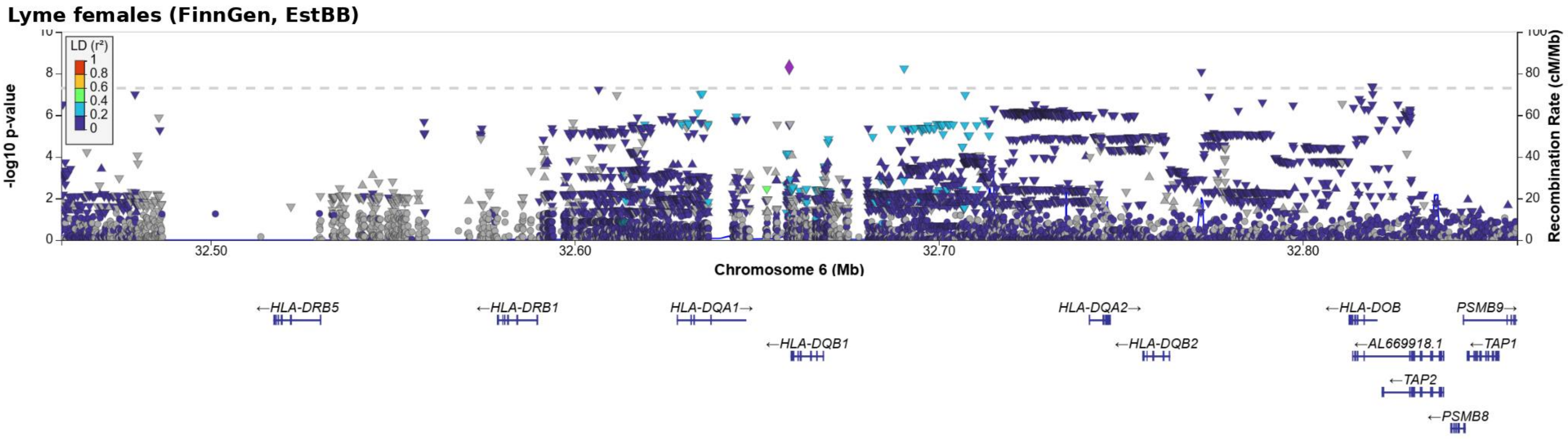


**Supplementary Figure S8.** Regional association plot for the HLA locus (chromosome 6) in the female-stratified Lyme disease GWAS meta-analysis. Points are colored according to linkage disequilibrium (LD, r²) with the lead variant. The dashed horizontal line indicates genome-wide significance ($P = 5 \times 10^{-8}$). The blue line denotes recombination rate, and gene annotations are shown below the association plot.

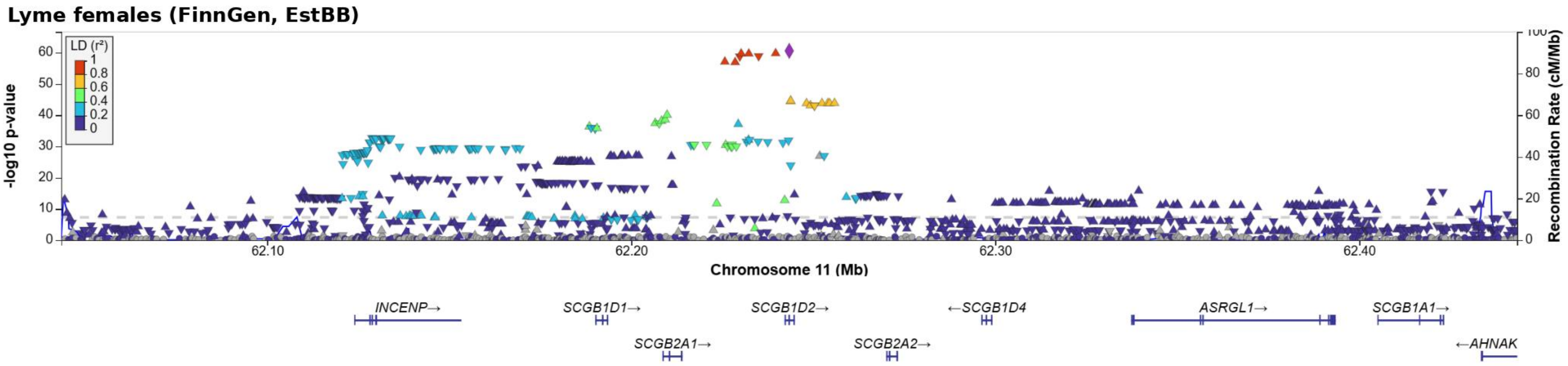


**Supplementary Figure S9.** Regional association plot for the *SCGB1D2* locus (chromosome 11) in the female-stratified Lyme disease GWAS meta-analysis. Points are colored according to linkage disequilibrium (LD, $r^2$) with the lead variant. The dashed horizontal line indicates genome-wide significance ($P = 5 \times 10^{-8}$). The blue line denotes recombination rate, and gene annotations are shown below the association plot.

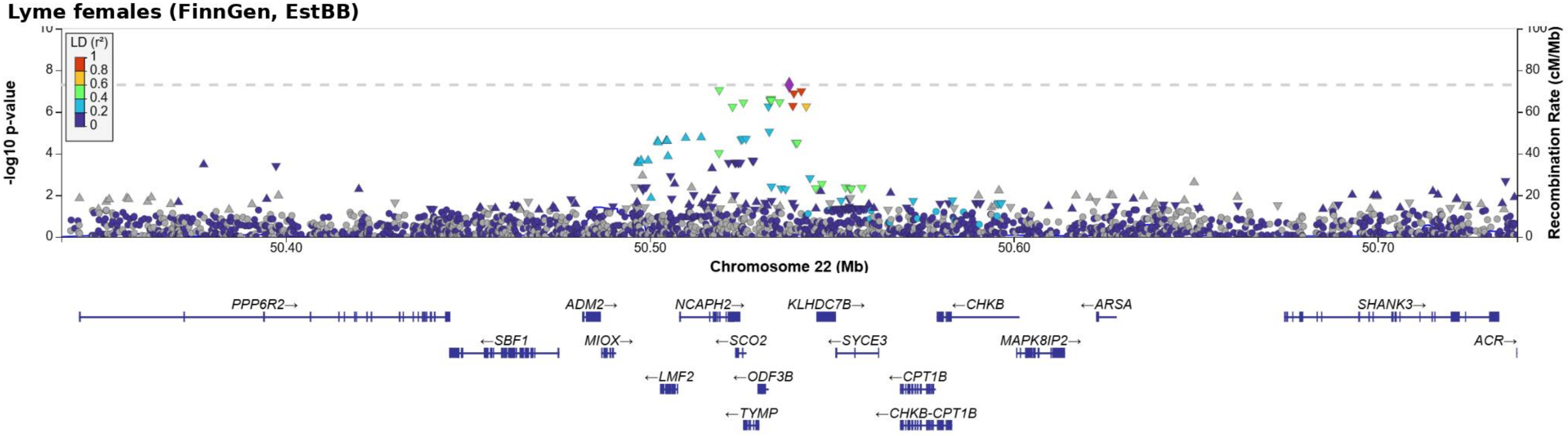


**Supplementary Figure S10.** Regional association plot for the *TYMP-CPT1B* locus (chromosome 22) in the female-stratified Lyme disease GWAS meta-analysis. Points are colored according to linkage disequilibrium (LD, r²) with the lead variant. The dashed horizontal line indicates genome-wide significance (P = 5 × 10⁻8). The blue line denotes recombination rate, and gene annotations are shown below the association plot.

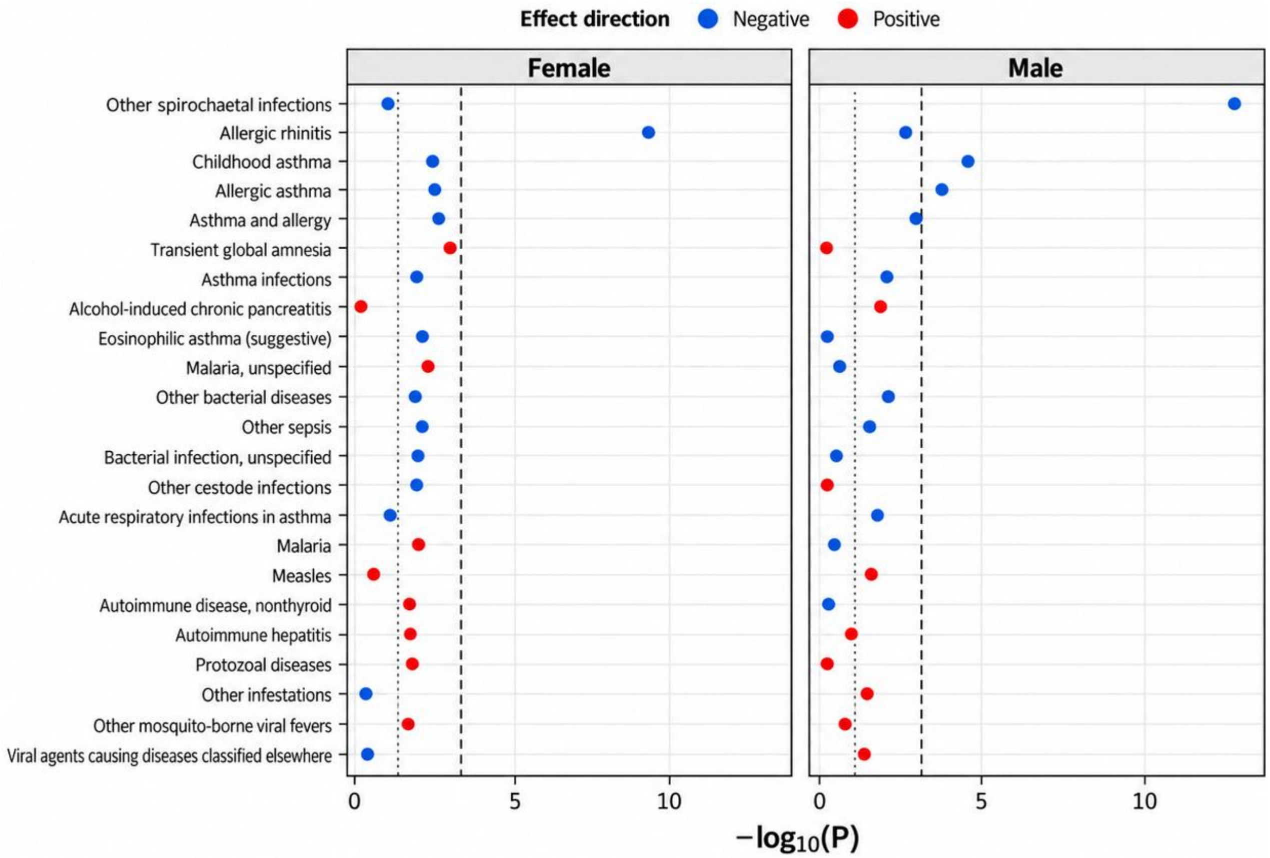


**Supplementary Figure S11.** Sex-stratified PheWAS of the *TLR1/6* lead variant rs148906978 in FinnGen. Phenotypes with nominal association (P < 0.05) in either females or males are shown in both panels. Points indicate $-\log_{10}(P)$, with color denoting the direction of the allelic effect. The dotted line indicates P = 0.05 and the dashed line the Bonferroni-corrected significance threshold (P = 0.00035). The "Other spirochaetal infections" endpoint includes Lyme disease and therefore does not represent an independent association with non-Lyme spirochaetal infections.
